# Defining a master curve of abdominal aortic aneurysm growth and its potential utility of clinical management

**DOI:** 10.1101/2020.10.28.20221101

**Authors:** Emrah Akkoyun, Hamidreza Gharahi, Sebastian T. Kwon, Byron A. Zambrano, Akshay Rao, Aybar C. Acar, Whal Lee, Seungik Baek

**Author notes:** Address for Correspondence: S. Baek, Ph.D., Department of Mechanical Engineering, Michigan State University, 2457 Engineering Building, East Lansing, MI 48824.

## Abstract

**Objective:** The maximum diameter measurement of an abdominal aortic aneurysm (AAA), which depends on orthogonal and axial cross-sections or maximally inscribed spheres within the AAA, plays a significant role in the clinical decision making process. This study aims to build a large dataset of morphological parameters from longitudinal CT scans and analyze their correlations. Furthermore, this work explores the existence of a “master curve” of AAA growth, and tests which parameters serve to enhance its predictability for clinical use.

**Methods:** 106 CT scan images from 25 Korean AAA patients were retrospectively obtained. We subsequently computed morphological parameters, growth rates, and pair-wise correlations, and attempted to enhance the predictability of the growth for high-risk aneurysms using non-linear curve fitting and least-square minimization.

**Results:** An exponential AAA growth model was fitted to the maximum spherical diameter, as the best representative of the growth among all parameters (r-square: 0.985) and correctly predicted to 74 of 79 scans based on a 95% confidence interval. AAA volume expansion rateswere highly correlated (r=0.80) with thrombus accumulation rates.

**Conclusions:** The exponential growth model using spherical diameter provides useful information about progression of aneurysm size and enables AAA growth rate extrapolation during a given surveillance period.

## Introduction

An abdominal aortic aneurysm (AAA) is characterized by a permanent dilation of the abdominal aorta (30 mm or more) [1, 2]. The decision to intervene is made based on AAA size measured by a maximum diameter (5.5 cm for men, 5.0 cm for women) or its growth rate (1 cm per year) [3, 4]. For small AAAs, long-term monitoring is recommended prior to any surgical intervention (open surgery or endovascular aortic repair (EVAR)). However, 10 – 24% of aneurysms below the intervention threshold (< 55mm) experience rupture as shown in some series [5, 6]. Therefore, the guideline for the non-surgical management of AAA reported that one of the unresolved issue was the development of better predictive tools for individual rupture risk including morphology based indicators. Additionally, a rapid expansion of AAAs is often associated with higher rupture risk [7–9], and it has long been suggested that the annual growth rate may play a role in prognosis, surgical planning, and patient management.

Although time-dependent geometrical analysis is a significant part of the clinical decision making process, quantification of the expansion rate remains ambiguous. Multiple studies have suggested that variability of AAA expansion rates is high, both over time in the same patient and among various patients [10, 11]. Furthermore, finding the natural growth pattern is difficult as the change of diameter is small and non-linear [12]. Studies have also reported that growth ratesare not constant; instead, periods of active rapid growth are followed by periods of non-activity [13, 14]. However, others have suggested a general AAA growth pattern in which an AAA expands over time with an increasing expansion rate as it gets larger [2, 9, 15–17].

Gharahi et al. [15] presented a method (“maximally inscribed spherical diameters”) based on largest spheres that fit within the AAA. Their analysis on a data set of 59 scans from 14 patients showed that the spherical diameter measurement provides the least variability, compared to the axial and orthogonal diameter measurements. In addition, they proposed that an exponential function fits the AAA growth pattern. These preliminary results call for a more extensive study on the AAA morphological growth patterns and the correlation between geometrical characteristics and their growth rates. Furthermore, this study investigates whether there exists a potential population-based pattern of AAA growth rate for a specific group of patients. This work will explore the idea of a “master curve” for AAA growth, and the parameters which best enhance its predictability for clinical use.

To this end, we retrospectively obtained longitudinal CT scan images of a Korean cohort, computed a total of 21 morphological parameters, and investigated their correlations to identify the best parameters to predict the AAA growth pattern.

### Patient and methods

#### Study Designs and Populations

118 computed tomography (CT) scans from 26 patients at the Seoul National University Hospital were used for this analysis. Patients were monitored and scanned at various time intervals between 3 to 56 months with a median interval of 11 months. Images were obtained using a CT scanner (Siemens Healthcare, Erlangen, Germany). The image in-plane resolution was 0.641 mm and the axial (z-axis) resolution of 1 mm. This study was subject to Internal Review Board approvals at Michigan State University and Seoul National University Hospital. Since our study does not involve identifiable human subjects, and only processes anonymized and archived CT scans, the need for ethical approval was waived by the Michigan State University Institutional Review Board (reference: IRB# 12-1041).

All AAAs with at least two CT scans and a time interval of at least 6 months were used for this study. This inclusion criterion was made in order to minimize the growth rate error. As a result, 106 CT scans from 25 patients (23 men and 2 women) were used and 21 different parameters describing the geometric properties of each scan were calculated. The mean age at time of first scan was 59 years old (55-84), with a 13month mean time between scans (6-56), and 4 scans per person (2-7).

#### Geometrical representation

From these CT images, AAA geometries were reconstructed using a biomedical software, Mimics® (Materialise, Leuven, Belgium), following the procedure previously described (Gharahi et al. [15]; Kwon et al. [18]). The centerline for each of these geometries was generated using the *maximally inscribed spheres* method. A series of slices perpendicular to the centerline (orthogonal planes), or to the Z-axis (axial planes), were made with a constant interval distance, such that the intersection of these planes with the AAA surface produced the cross-section required to measure the orthogonal or axial parameters.

The definitions of all geometrical measurements, which were classified as either primary or secondary parameters, are summarized in **Table 1**. These parameters reflect the properties of the aneurysm at the time of the scan. In addition, details of maximum diameter measurements are described by Gharahi et al. [15].

**Table I.** The definitions of geometrical measurements

|  | Parameter | Description |
| --- | --- | --- |
| **Primary** | $MAX_{DIA\_S}$ | the maximum inscribed spherical diameter |
| | $MAX_{DIA\_A}, MAX_{DIA\_O}$ | the maximum axial and orthogonal diameter |
| | $MAX_{PER\_A}, MAX_{PER\_O}$ | the maximum perimeter on axial and orthogonal planes |
| | $DIA_{PER\_A}, DIA_{PER\_O}$ | the perimeter on axial and orthogonal planes at the maximum spherical diameter |
| | $VOL_{AAA}$ | the total volume of the aneurysm |
| **Secondary** | $VOL_{ILT}$ | the total volume of the thrombus |
| | $VOL_{LUMEN}$ | the total volume of the lumen |
| | $MIN_{DIA\_A}, MIN_{DIA\_O}$ | the minimum diameter on axial and orthogonal planes |
| | $MAX_{ECC\_A}, MAX_{ECC\_O}$ | the maximum eccentricity on axial and orthogonal planes |
| | $DIA_{ECC\_A}, DIA_{ECC\_O}$ | the eccentricity on axial and orthogonal planes at the maximum spherical diameter |
| | $MAX_{ILT}$ | the maximum thrombus thickness |
| | $A_{ILT}$ | the fraction of AAA surface area covered by ILT content |
| | $MDIA_{TORT\_DISP\_A}, MDIA_{TORT\_DISP\_O}$ | the displacement of the AAA centerline from the line joining the first and last points of the AAA centerline at the maximum diameter cross section, on axial and orthogonal planes |
| | $TORT\_CL$ | the ratio of the total centerline length to the length of the line joining the first and last point |

Eccentricity was defined as the ratio of maximum to minimum diameter and was calculated for both orthogonal and axial planes. Tortuosity of the centerline was calculated as the ratio of the total centerline length to the length of the line joining the first and last point. Finally, perimeter was found by measuring the length of the line forming the boundary of the aneurysm shape in the cross-sectional plane.

Volume measurements, denoted as VOL, include lumen volume, total AAA volume, and calculated intraluminal thrombus (ILT) volume (subtracting the total lumen volume from the total AAA volume). The global maximum and minimum of a local measurement in AAA geometry were denoted by MAX and MIN, respectively.

#### Growth rates and their correlation analysis

The growth rate of each AAA patient at a given time was computed by considering changes in parameters between two consecutive scans. The rate values for all measurements were calculated based on the assumption that there is an exponential change of the measurement over time. The rate of a parameter *g*(.), introduced in [16], is computed for all geometrical measurements by using the following equations

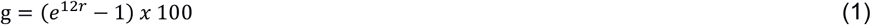

where *r* is the logarithmic growth factor measured by

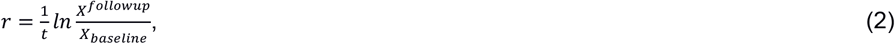

*X*(.) denotes the quantity of the geometrical measurements and *t* is the time interval between consecutive scans in months. For example, *g*MAX_DIA_S_ was used to define the growth rate of maximum spherical diameter, where the maximum spherical diameters can be compared at baseline and follow-up. Pearson correlation coefficients were used for correlation analysis of different growth rates. We consider correlation ranges of 1.00–0.90, 0.90–0.75, 0.75–0.50, 0.50–0.25, 0.25–0.0 as very high, high, moderate, weak, and no correlation, respectively.

#### Exponential AAA growth model and the growth prediction

All geometric measurements were analyzed to evaluate potential capability of growth prediction. As proposed by Martufi et al. [16] the growth rate was expressed using an exponential function over the time. The growth curve is defined as an exponential function

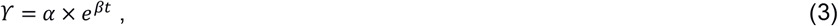

where *γ* is the measurement and *α* and *β* are parameters of the growth curve. The input time *t* is the shared time axis for all the patients. Since the AAA stage of the patients at the time of first scan was not the same, the time of the scan must be shifted in the shared time axis. Exponential AAA growth model is based on the assumption that the individual growth patterns, the parameters of each growth curve (*α* and *β*) are identical to the master curve pattern.For this purpose, an initial growth curve is fitted (*α* and *β*) to one patient. Subsequently, the time of the first scan 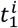 is estimated using the method of least squares so that the measurement set best fits the common growth curve, where superscript *i* denotes patient data sets. These two steps are repeated, updating 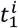 at each iteration until convergence(i.e. total amount of error met the convergence criterion or no longer decreases) is achieved. The psuedocode of the iterative algorithm is given in **Appendix I** (**Figure A1**). Finally, the growth curve, named the *master curve*, was obtained using fminsearch, a built-in function of MATLAB software to find a local minimum for unconstrained nonlinear optimizationbased on the Nelder-Mead simplex algorithm.

The least fluctuations and narrowest range in measurements contributes to the prediction strength of the master curve, obtained to better summarize the growth and to evaluate the prediction accuracy for each measurement. The coefficient of determination, denoted by r-square, was utilized to measure how well growth was predicted [19], such that the minimum proportion of the total variance of outcomes explained by the model was selected as the most representative of the growth curve. The curve was log transformed from non-linear to linear and evaluated by r-square.

## Results

All geometrical measurements describing the geometric properties of each scan were calculated. First, these measurements were analyzed in terms of their correlations regardless of time interval between consecutive scans. Second, the growth rate of each measurement was computed by considering changes in parameters between two consecutive scans. Finally, growth curves, namely master curves, were constructed for these measurements to evaluate prediction accuracy. All these results were sequentially given under proper titles below.

### Maximum measurements for correlation analysis

A total of 21 measurements were analyzed in terms of their correlations for each observation and are summarized in **Table 2**. The correlation study showed that AAA volume is highly correlated with diameter, regardless of method used to calculate the maximum diameter (spherical (r=0.89), axial (r=0.91) or orthogonal (r=0.92)). Similarly, very high correlations were found between the diameters and perimeters regardless of methods used (r>0.92). All primary parameters (r=0.69 and 0.77) are mildly correlated with ILT volume. However, the secondary parameters are significantly less correlated with the primary parameters, except ILT volume. For instance, eccentricity (r=0.60) and tortuosity (r=0.55) are moderately correlated with AAA volume; only maximum ILT thickness (r=0.52-0.59) is moderately correlated with maximum diameter measurements. **Table II**. Correlations of geometrical parameters on AAA actual measurements

**Table II.** Correlations of geometrical parameters on AAA actual measurements

|  | **Parameters** | **VOL<sub>AAA</sub>** | **VOL<sub>ILT</sub>** | **MAX<sub>DIA_S</sub>** | **MAX<sub>DIA_O</sub>** | **MAX<sub>DIA_A</sub>** | **MAX<sub>PER_O</sub>** | **MAX<sub>PER_A</sub>** |
| --- | --- | --- | --- | --- | --- | --- | --- | --- |
| **1°** | VOL<sub>AAA</sub> | 1 |  |  |  |  |  |  |
|  | MAX<sub>DIA_S</sub> | **0.89** | 0.69 | 1 |  |  |  |  |
|  | MAX<sub>DIA_O</sub> | **0.92** | 0.72 | **0.92** | 1 |  |  |  |
|  | MAX<sub>DIA_A</sub> | **0.91** | **0.77** | **0.94** | **0.95** | 1 |  |  |
|  | MAX<sub>PER_O</sub> | **0.93** | 0.74 | **0.96** | **0.96** | **0.96** | 1 |  |
|  | MAX<sub>PER_A</sub> | **0.92** | **0.75** | **0.93** | **0.92** | **0.94** | **0.94** | 1 |
| **2°** | VOL<sub>ILT</sub> | **0.78** | 1 |  |  |  |  |  |
|  | MAX<sub>ECC_O</sub> | 0.60 | 0.08 | 0.22 | 0.47 | 0.33 | 0.28 | 0.30 |
|  | TORT_CL | 0.55 | -0.16 | 0.04 | 0.07 | 0.01 | 0.03 | 0.03 |
|  | MAX<sub>ILT</sub> | 0.46 | **0.81** | 0.53 | 0.52 | 0.59 | 0.56 | 0.59 |
|  | A<sub>ILT</sub> | 0.35 | 0.70 | 0.39 | 0.42 | 0.47 | 0.42 | 0.49 |

### Growth rates and correlation analysis

Growth rates of the maximum spherical (median 6.04%/year, IQR 5.66%/year), orthogonal (median 6.47%/year, IQR 7.14%/year), and axial (median 5.75%/year, IQR 5.95%/year) diameters, as well as aneurysm volumes (median 13.44%/year, IQR 15.12%/year) are summarized via Box and Whisker plots in **APPENDIX I** (**Figure A2**).

The normality of the diameters and the aneurysm volume growth were analyzed using the Shapiro-Wilk test, and the result confirmed the normality of the diameters (spherical p=0.87, axial p=0.27 and orthogonal p=0.53) but not the volume (p=0.01). A Mann Whitney U test indicated that the diameters were not significantly different from each other (p>0.77). However, the growth rates of these diameters were significantly different from that of the aneurysm volume growth rate (p<0.01).

In addition, the growth rate for each measurement was calculated in a non-linear fashion and their pairwise correlation was analyzed in **Table 3**. AAA volume expansion rates are highly correlated with axial diameter growth rates (r=0.80), and moderately correlated with the spherical (r=0.61) and orthogonal (r=0.72) diameter growth rates. Orthogonal and axial diameter growth rates have a strong correlation with each other (r=0.78), whereas these two rates do not show a strong correlation with spherical diameter growth rates (r=0.55 and 0.67, respectively).

**Table III.**
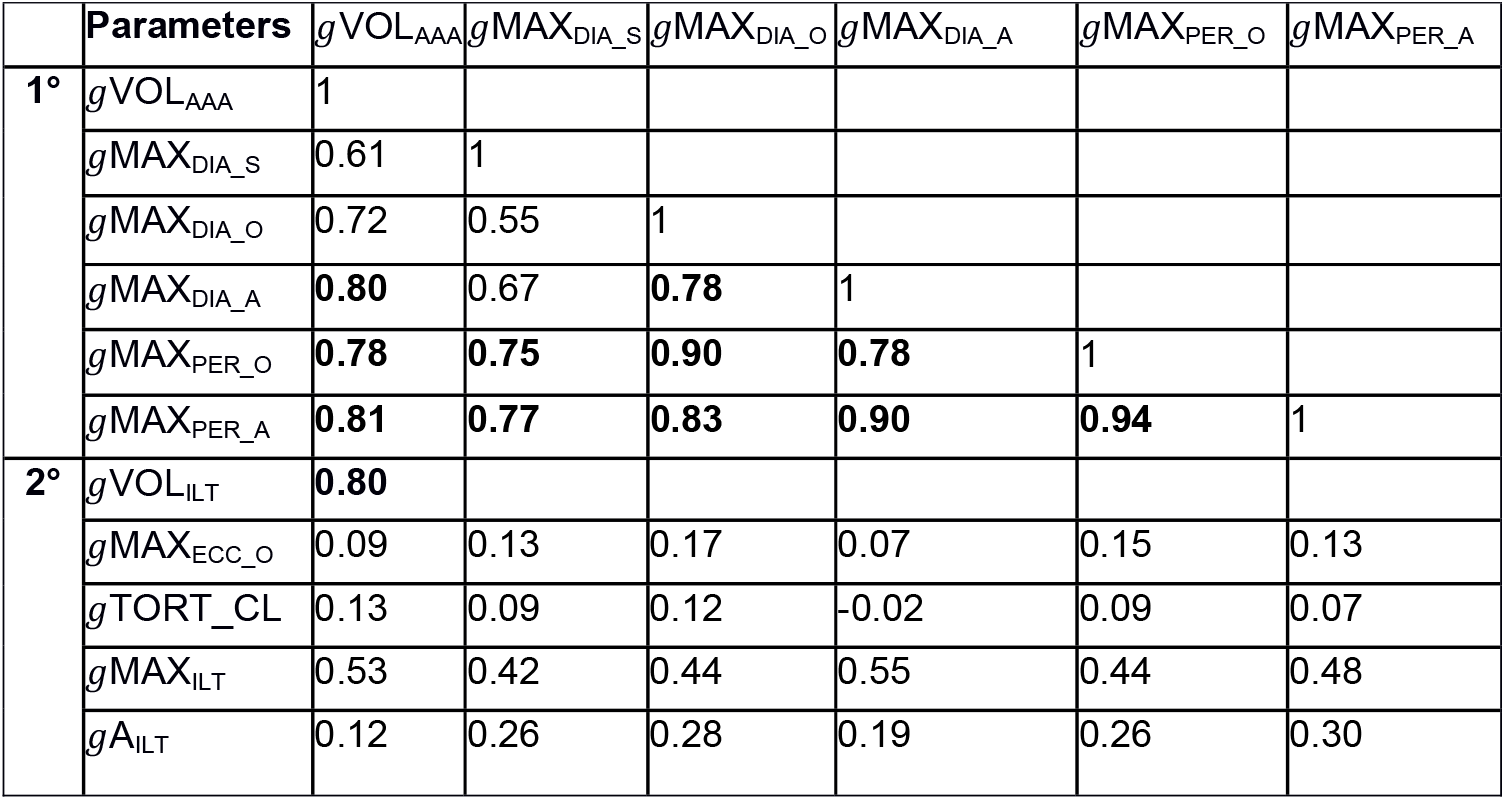
Correlations of geometrical parameter rates of change on AAA measurements using non-linear growth model

AAA volume expansion rate is highly correlated (r=0.80) with thrombus accumulation rate. However, the aneurysm volume expansion rate is not highly correlated with the maximum ILT thickness (r=0.53) nor with the lumen volume (r=0.45) rates. Other secondary parameters such as eccentricity, tortuosity, ILT thickness and area fraction rates are not highly correlated with AAA volumetric expansion rates (r=0.09, 0.13, 0.53 and 0.12 respectively). Additionally, eccentricity and tortuosity parameters are not correlated with any of the primary parameters (r<0.17).

### Growth curve of the geometric measurements

Growth curves were constructed for the measurements in order to find one that could predict aneurysmal growth. **Figure 1** compares the master curves obtained for maximum spherical diameter (left) and orthogonal diameter (right). The best three representatives of the master curves were selected based on their r-square scores. The spherical diameter (MAX_DIA_S_) was found to be the best growth representative (r-square: 0.985) and the three next-best representatives were MAX_PER_A_ (0.977), DIA_PER_A_ (0.972), andMAX_DIA_O_ (0.970). **Figure 2** shows the prediction of AAA growth based on the master curve and the histogram of error prediction, based on spherical diameter.

**Figure 1.**
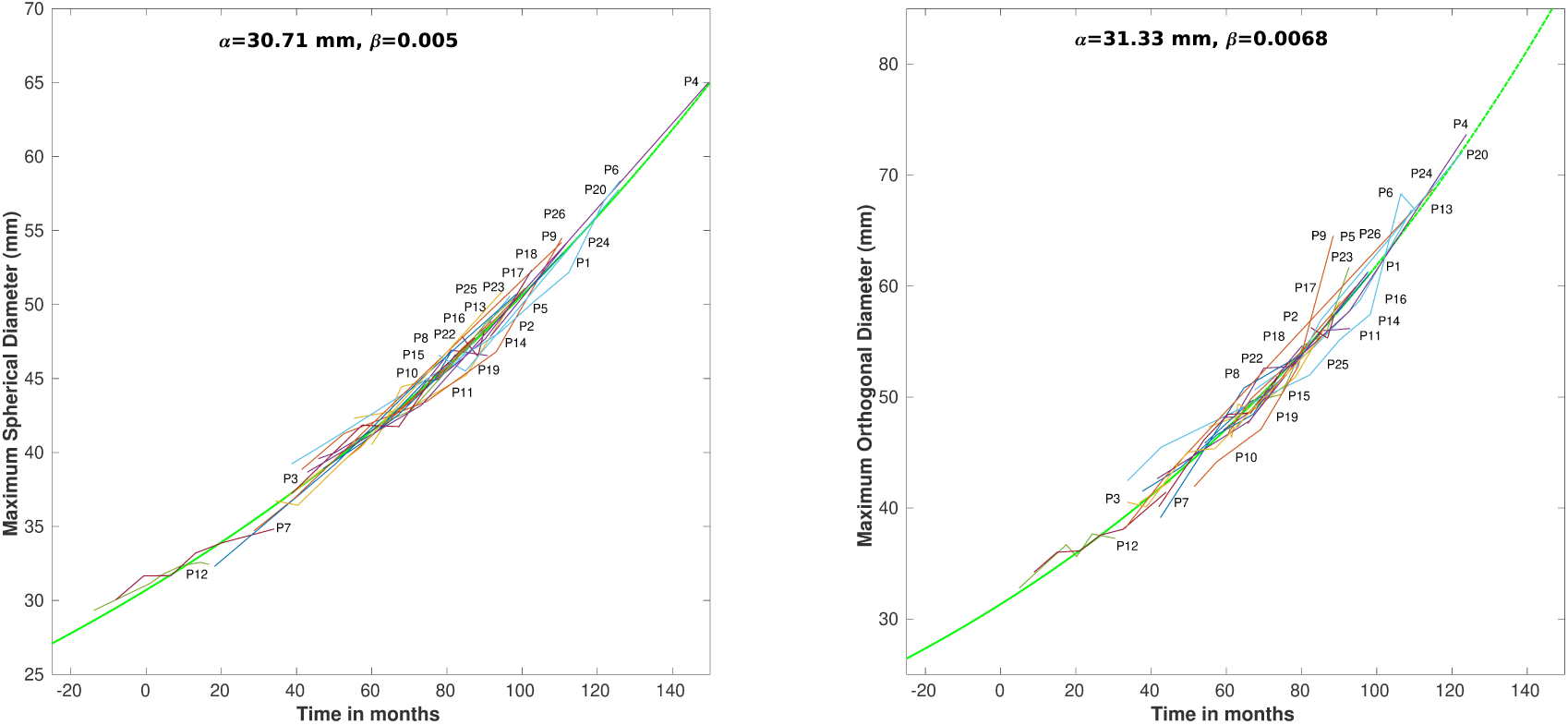
The exponential functions of maximum spherical diameter (left) and maximum orthogonal diameter (right). The spherical diameter is the best representative of AAA growth

**Figure 2.**
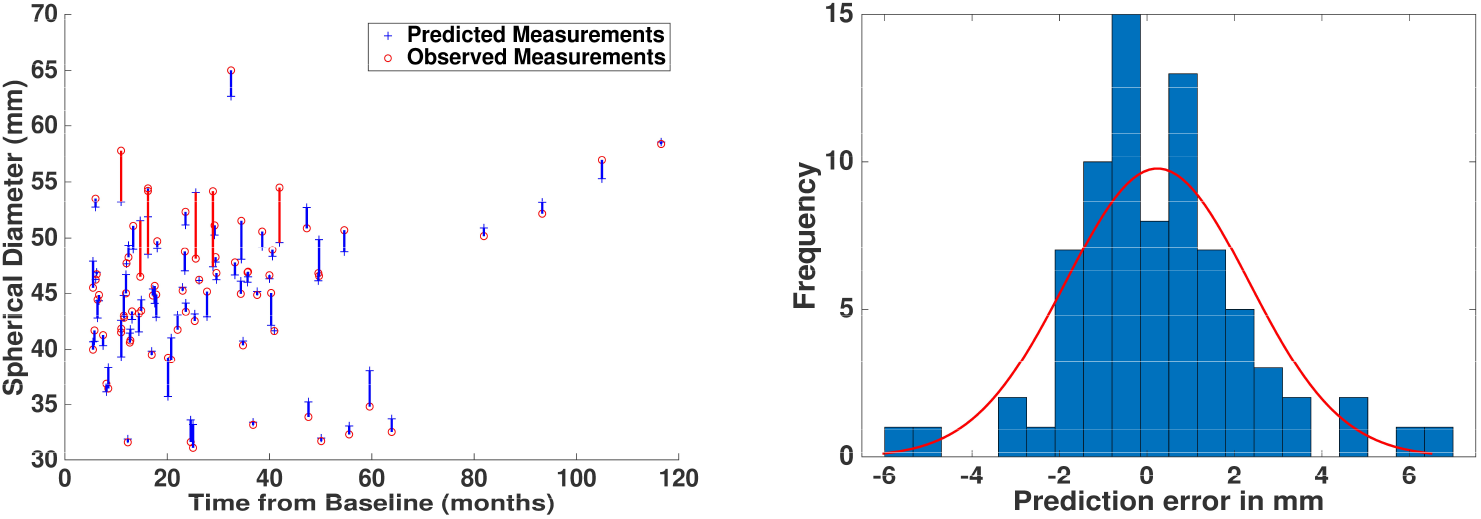
The prediction of AAA growth based on the master curve (spherical diameter). a) The actual and predicted values are plotted with respect to time from baseline. b) The histogram of prediction errors and estimation of normal distribution parameters

Using the master curve function derived (mean=0.24 mm, sigma=2.10 mm), the spherical diameter was correctly predicted in 74 of 79 scans (visualized by blue lines Fig. 3a), based on a 95% confidence interval. Similarly, the prediction capabilities of other diameter measurements were found to have averages of 0.52 mm and 0.08 mm, and standard deviations of and 3.23 mm and 2.68 mm, for orthogonal and axial, respectively.

## Discussion

Previous studies suggested that orthogonal diameter yields the measure closest to real AAA size, and is superior to axial diameter, which tends to overestimate the diameters [20, 21]. The orthogonal diameter measurement method, however, can be dependent on the construction of the centerline, which can be highly variable. For instance, an error of 5°in determining the orthogonal plane might lead to 15 mm of miscalculation in measuring maximum diameters [15]. To address this issue, this study utilized a method to reduce variability which semi-automatically generates the centerline using maximally inscribed spheres.

The guideline for clinical AAA management based on single maximum diameter criterion has been challenged [5, 6, 9, 16], with more studies proposing that the growth rate is associated with AAA rupture [7, 9]. An augmented criterion, the maximum diameter > 5.5 cm or annual growth rate >1 cm/year, has been proposed for surgical intervention [3, 4]. There is, however, scarcity of morphological studies using longitudinal CT scan images. Therefore, this work aimed to construct a larger database of morphological parameters and to enhance the predictability AAA growth for high-risk aneurysms.

Previous studies [16, 20] proposed that the aneurysmal volume measurement served to better predict the development of AAA and rupture risk than the maximum diameter measurement. One of our main findings, however, is that the total volume is highly correlated with all primary parameters (maximum diameters, perimeter).

Meanwhile, there has been increasing evidence that growth rate is important for predicting high rupture risk [7, 9]. AAA volume expansion rates are only mildly correlated with the spherical (r=0.68) and orthogonal growth rates (r=0.67). Furthermore, spherical diameter growth rates do not show strong correlations with axial and orthogonal diameter growth rates (r=0.55 and 0.72, respectively). Although the utilities of different maximum diameter measurements and their rates were not fully tested, Gharahi et al. [15], suggested that different diameter measurements may serve different purposes. In particular, the axial diameter measurement is conveniently determined without finding a centerline [20], orthogonal diameter is important for representing the actual size and assessing the rupture potential [20, 21], and the spherical diameter is potentially suitable for predicting the AAA growth.

AAA volume expansion rate is highly correlated (r=0.80) with thrombus accumulation rate.Zambrano et al [22] demonstrated that ILT was initially formed at the region of aorta where low wall shear stress was observed and its accumulation rate was associated with the aneurysm’s expansion rate. Similarly, Parr et al. [23] found that the aneurysm volume was correlated with thrombus volume and diameter. The secondary parameters such as eccentricity, tortuosity, area fraction covered by ILT, ILT thickness, and lumen volume present no strong correlation with AAA volume expansion in terms of geometrical (static) (r < 0.45) and rate measurements (r < 0.50). These parameters, however, might be still important for the assessment of rupture risk. In fact, previous studies reported that some parameters such as asymmetry and tortuosity [24], and ratio of ILT to AAA volume [25] are associated with rupture risk.

We used an exponential function [2, 16, 17] for modeling AAA growth since AAAs of 3-3.9 cm size expand slowly (a mean growth rate of 2.84 mm/year) compared to AAAs of 4-4.9 cm size (a mean growth rate of 3.66 mm/year). Similarly, previous studies reported growth rates of 1.1-7 mm/year for AAAs with 3-3.9 cm initial diameter, in contrast to the growth rate of 3-6.9 mm/year for AAA with 4-4.9 cm [26]. In addition to the growth rate studies, we analyzed the outcomes according to the expansion pattern. It was observed that an AAA with a 3 cm diameter will need surgical repair within the first 7 years of the scan. However, an AAA with a 4 cm diameter will reach 5.5 cm in the first 3 years. The UK SAT demonstrated that less than 20% of patients with 3-3.9 cm AAA would need surgical repair in the first 5 years of follow-up [12, 27]. In the ADAM study, 27% of 4-5.5 cm AAA undergo surgical intervention in the first 2 years of follow-up [28].The recent AAA guideline (2019) recommended safe surveillance intervals such as every three years for aneurysms 3–3.9 cm in diameter, annually for aneurysms 4.0– 4.9 cm, and every 3–6 month for aneurysms ≥5.0 cm [3]. Therefore, the growth pattern computed in this follow-up study are consistent with those reported in the literature.

Most of all, this study found the spherical diameter as the best representative of the growth curve (r-square: 0.985) with a significantly higher prediction strength compared to other diameter measurements. Moreover, the maximally inscribed spheres method, minimizes the variability of the geometrical surface, and as a result, it leads to the least fluctuations and narrowest range in measurements [15].

We examined the utility of master curves and their prediction capabilities in terms of different geometrical parameters. The proposed model better predicts the growth of AAAs due to adoption of an exponential growth function, rather than a traditional linear model, and systematically consider the effects of 21 geometric measurements (i.e. independent variables) on the growth rate.Among all the parameters, the master curve of spherical diameter performed best, predicting the diameter within 0.42 mm in 95% of all scans. In addition, we observed that the master curve using the spherical diameter resulted in the smallest prediction error (sigma=2.10 mm), while those of orthogonal and axial diameter resulted in larger errors (sigma = 2.68 mm and 3.22 mm). Therefore, we propose that a master curve for spherical diameter may be used as a clinical tool that gives insight about the future of aneurysm growth. This predictive tool can be used for planning for follow-up scans and surgical interventions.

### Limitations

Although defining the master curve has a potential utility for predicting the aneurysm growth rate and, hence for clinical management, there are clear limitations. First of all, the master curve, established in this study, was based on a purely heuristic approach. Particularly, this study assumed that individual growth patterns are identical to the representative growth pattern, while the maximum diameters of AAA patients at the time of first scan were not identical to other patients. Despite the lack of understanding of the exactbiochemical mechanisms, various data-driven or feature-based apporaches have proven useful for medical application [16, 20, 26];this study might provide a new utilty for the accurate prediction of AAA growth rate. Second, decision-making related to clinical management for AAA patients is complicated because information of impending AAAs prior to rupture is rarely available or surrogates, for example, AAAsof high rupture risk that is required for immediate intervention can be used [3]. This study does not use ruptured CT scans, and the direct rupture risk assessment is beyond the scope of this study. Third, the AAA growth curve modelled here is only used for the assessment of the likelihood of an AAA rupture according to the maximum diameter protocols [2],but other factors such as the patient’s age, presence of coexistent peripheral artery disease, peripheral aneurysm and whether AAAs are asymptomatic/symptomatic may be important to consider when determining when to proceed with elective AAA repair [3,4].Lastly, this study only analyzed a subset of Korean patients from a single institution, thus the results may not be extrapolatable. Particularly, these findings of the master curve for the spherical diameters could not be compared to other results in literature. Regardless of these limitations, this study provides valuable information about aneurysm evolution using various geometrical measurements and offers an acceptable growth model for development of an improved surveillance program.

## Conclusions

The exponential growth model was constructed using various diameter measurements, and spherical diameter was found to be the best representative of growth. This measure provides useful information about the evolution of aneurysm size and may be clinically helpful. Nevertheless, the different diameter measurement methods may have their own capabilities and shortcomings, which lead to different interpretations of aneurysm growth.

## Data Availability

Data referred in the manuscript is not publicly available.

## Appendix I

**Figure A1.**
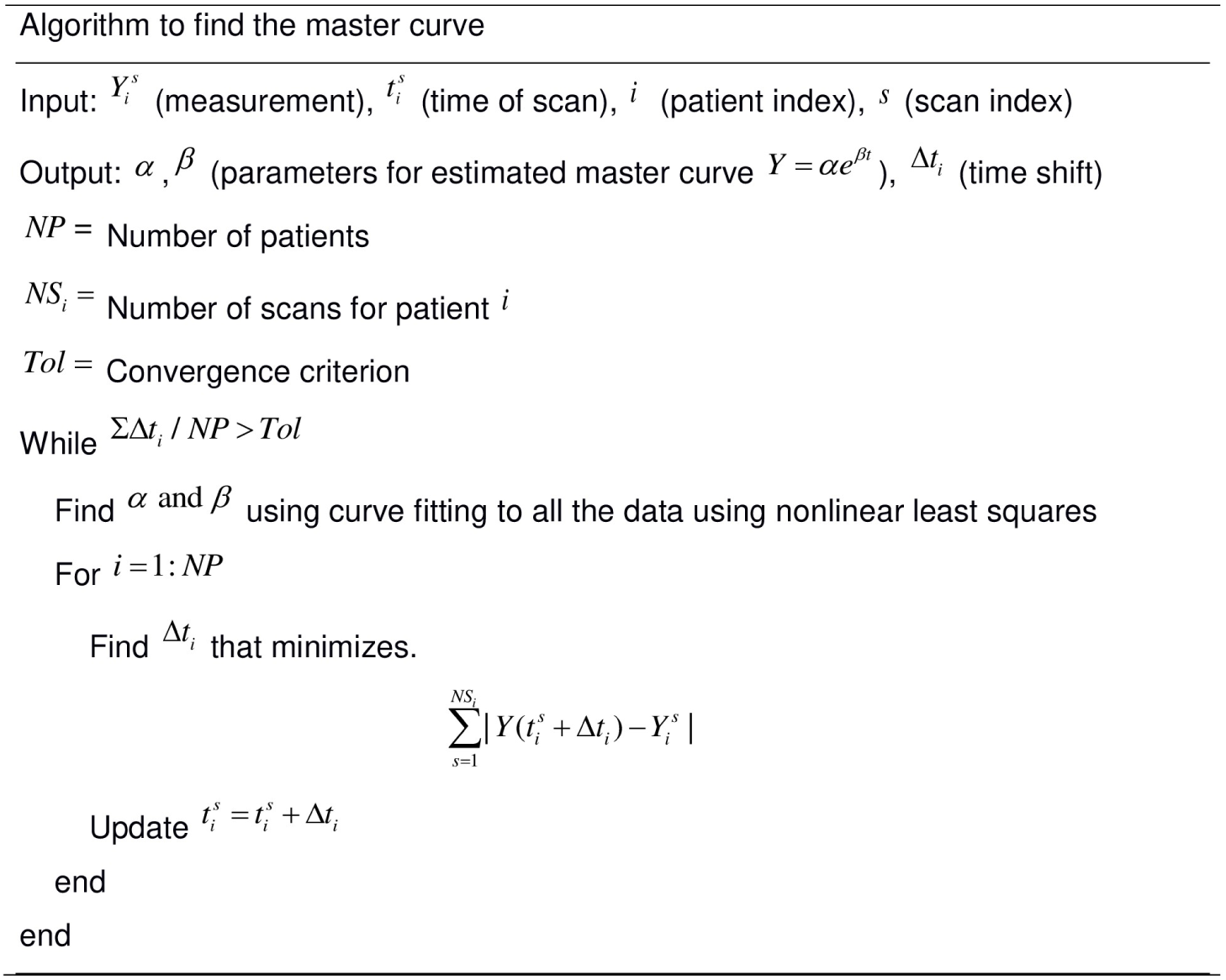
The iterative algorithm to find the master curve in Eq.3

**Figure A2.**
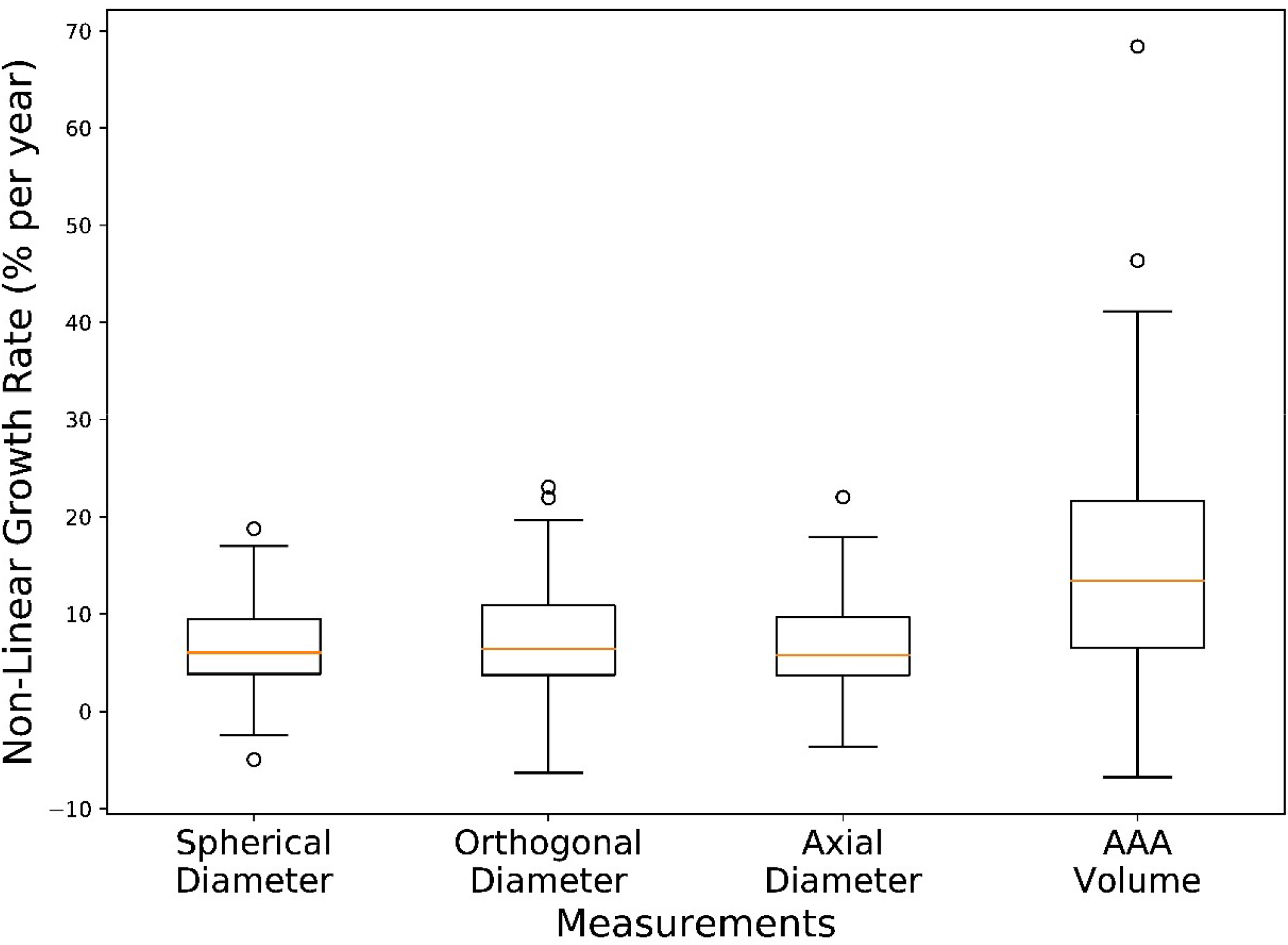
Box and whisker plots for the growth rate of the diameters and aneurysm volume using non-linear model

